# MassMark: A Highly Scalable Multiplex NGS-based Method for High-Throughput, Accurate and Sensitive Detection of SARS-CoV-2 for Mass Testing

**DOI:** 10.1101/2021.01.08.20249017

**Authors:** Kao Chin Ngeow, Chao Xie, Alvin Kuo Jing Teo, Li Yang Hsu, Min Han Tan, Yukti Choudhury

## Abstract

Mass testing has been proposed as a strategy to address and contain the severe acute respiratory syndrome coronavirus 2 (SARS-CoV-2) pandemic^1,2^. We have developed MassMark, a novel and highly scalable multiplex method that employs next generation sequencing for high-throughput, accurate and sensitive detection of SARS-CoV-2, while minimizing handling complexity and resources by utilizing a serial pooling strategy to accommodate over 9,000 samples per workflow. Analytical validation showed that MassMark was able to detect SARS-CoV-2 RNA down to a level of 100 copies per reaction. We evaluated the clinical performance of MassMark in a simulated screening testing with 22 characterized samples from three different sources (nasopharyngeal swabs, nasal swabs and saliva), comprising of 12 SARS-CoV-2 positive samples with mid to late Ct values (range: 22.98-32.72) and 10 negative samples. There was one false negative and no false positives, giving an overall sensitivity and specificity of 91.67% and 100% respectively, when compared against an optimized RT-PCR test with a target size within 70 bp (CDC 2019-nCoV Real-Time RT-PCR Diagnostic Panel^3^).

## Introduction

Since the first human cases of coronavirus disease 2019 (COVID-19) were reported in December 2019, the number of SARS-CoV-2 infections have continued to grow at an exponential rate, with over 79.2 million confirmed cases and over 1.7 million deaths reported worldwide as of 27 December 2020^4^. Furthermore, due to incomplete surveillance and imperfect diagnostic accuracy, the confirmed case numbers are likely to be a significant underestimate of the true number of SARS-CoV-2 infections^5–7^. While many countries were able to moderate the spread of the virus via nonpharmaceutical interventions such as lockdowns and travel restrictions^8^, the subsequent resurgence of cases in some areas^9^ demonstrates the necessity of constant and vigilant surveillance to control subsequent waves of SARS-CoV-2 outbreaks.

Though recent breakthroughs around SARS-CoV-2 vaccine development have been highly encouraging, with efficacy rates of 70-95% observed in Phase 3 clinical trials for three separate SARS-CoV-2 vaccine candidates^10–12^, manufacturing and distribution bottlenecks are likely to pose significant challenges to widespread SARS-CoV-2 vaccination coverage and limit its effectiveness in controlling the pandemic^13^. The number of total COVID-19 deaths worldwide is projected to be over three million by April 2021, even after taking into account the effect of vaccine rollouts^14^. Until effective SARS-CoV-2 vaccines are made broadly accessible and widely deployed, mass testing and other mitigation strategies will continue to play important roles.

Mass testing has been proposed as a strategy to contain and break the cycle of SARS-CoV-2 transmission^1,2^ while minimizing the need for drastic restrictive measures such as lockdowns, for which extended implementation is impractical due to severe, negative social and economic impacts^15^. Population-wide mass surveillance will also allow the detection of asymptomatic cases, estimated to be approximately 20% of total cases^16,17^, which form a contagious pool that contributes to viral transmission^18,19^ but are difficult to identify with existing symptom-based detection protocols.

While SARS-CoV-2 diagnostic testing capacity has increased considerably worldwide since the start of the pandemic, there is still a vast and significant gap between actual testing capacity and benchmark numbers needed to suppress the spread of the SARS-CoV-2 virus^1,20^. Inadequate testing capacity has led to testing backlogs and increased turnaround times^21^, hampering efforts to curtail infections^22^. Other than the gold standard RT-PCR method, alternative testing modalities will need to be introduced to overcome logistical bottlenecks and increase testing throughput in order for mass testing to become a feasible reality. Here, we describe MassMark, a novel method to scale-up SARS-CoV-2 diagnostic testing capacity via the use of massively parallel next-generation sequencing (NGS).

Like other proposed NGS-based approaches for SARS-CoV-2 diagnostics^23–27^, MassMark utilizes molecular barcodes to identify positive cases by incorporating a unique, predetermined combination of index sequences to amplify the viral RNA present in a positive sample, while samples negative for SARS-CoV-2 do not get amplified. The molecular barcode tagging is achieved in two steps with serial reduction in handling complexity, using only two sets of 96 unique index sequences for over 9000 samples. The use of molecular barcodes allows thousands of patient samples to be sequenced in parallel in a single NGS run, after which demultiplexing is performed via the barcode sequences to retrieve the identity of samples that are positive for SARS-CoV-2. We demonstrate that MassMark can be used for highly sensitive and specific detection of extracted SARS-CoV-2 RNA from nasopharyngeal swabs, nasal swabs and saliva.

## Results

The MassMark method (Fig 1A) consists of two serial rounds of barcoding with uniquely indexed primers and sample pooling. In the first step, extracted viral RNA is specifically converted to complementary DNA (cDNA) via reverse transcription (RT) at three target regions of the SARS-CoV-2 virus with three separate barcoding RT primers. Each barcoding RT primer consists of a region of homology to its specific target region of SARS CoV-2 and a predetermined 10-bp index sequence corresponding to the sample well position in a 96-well plate. For each well, the three RT primers carry the same 10-bp index sequence. During the RT step, individual samples are labelled with a unique sample index (well barcode). A sample that is negative for SARS-CoV-2 will fail to incorporate the corresponding index sequences. Products of cDNA synthesis from each 96-well plate are then combined, concentrated and purified together in a single pool.

**Figure 1:**
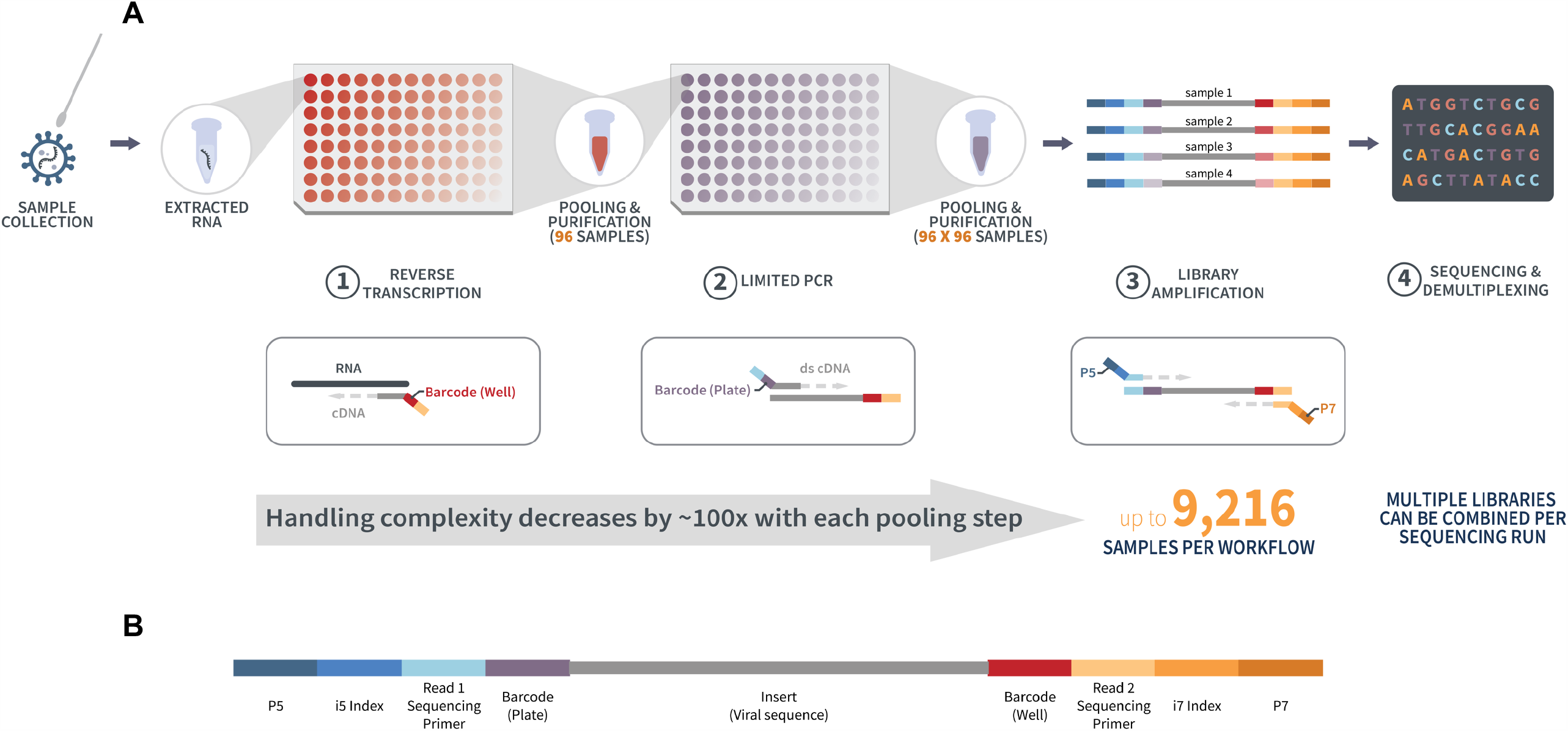
**(A)** Overview of MassMark workflow. 1) Extracted RNA undergoes reverse transcription to generate cDNA tagged with a barcode corresponding to the sample well position in the plate, following which up to 96 samples are pooled per plate. Only one of up to 96 plates for one workflow is represented in the figure. The configuration of sample barcodes is the same on each plate. 2) Single-stranded cDNA undergoes a limited, 1-cycle PCR to generate double-stranded cDNA. A second barcode representing the plate identity is added in this PCR. Samples from up to 96 plates are then pooled together. 3) The double-stranded cDNA pool undergoes PCR amplification, during which Illumina sequencing adapters and library indexes are attached. 4) The final library representing up to 9,216 samples is sequenced, and samples are demultiplexed according to their unique combination of well and plate barcodes. Multiple libraries can be sequenced in the same run and demultiplexed according to their library indexes. **(B)** Structure of final dually barcoded product to be sequenced.

Each cDNA pool originating from a single 96-well plate is then subjected to a limited PCR cycle with a second set of three barcoding primers, each consisting of a region of homology to its specific target region of SARS CoV-2 and a predetermined 10-bp index sequence corresponding to the plate identity. The limited PCR converts the products of the first step of cDNA synthesis to double-stranded DNA products. Importantly, the limited PCR step additionally labels each individual sample with a second index (plate barcode) common to all samples originating from the same RT plate. Together with the first sample index (well barcode) introduced during RT, the addition of the second sample index means that each sample will be in possession of a unique dual-index combination that allows for subsequent demultiplexing. Products of limited PCR (up to 96) are combined, concentrated and purified together, resulting in a single pool of pools that represents SARS-CoV-2 amplicon sequences from up to 96×96 samples.

This double-stranded 96×96 DNA pool then undergoes a final PCR amplification step to introduce adapters compatible with Illumina sequencing, and, optionally, to attach additional library indexes to allow multiple 96×96 pools to be multiplexed in the same sequencing run (Fig 1B). The final amplified library is sequenced, following which sequencing reads corresponding to each amplicon are identified by their expected SARS-CoV-2 sequence and are assigned to a starting sample by the identifiable, unique combination of well and plate barcodes associated with each amplicon that was finally represented in the sequencing library. An external spike-in control RNA is included in every well as a process control.

### Analytical validation

We verified that all three amplicons A, B and C were amplified in positive control samples comprised of synthetic SARS-CoV-2 RNA, diluted to known copy numbers with nuclease-free water. We observed a linear relationship between sequencing read counts and the amount of RNA input from approximately 100 to 50,000 copies per reaction, for all three amplicons (Fig 2). To determine the analytical limit of detection for MassMark, we repeated the protocol on 96 contrived samples comprising of 0 copies (n=56), 100 copies (n=20) and 250 copies (n=20) of synthetic SARS-CoV-2 RNA (Fig 3). At 100 input copies of SARS-CoV-2, reads above noise level (i.e. above read counts for 0-copy samples, in theory expected to be zero) corresponding to Amplicons A, B and C were detected in 17 (85%), 18 (90%) and 19 (85%) samples respectively, out of a total of 20 samples. At 250 input copies, reads above noise level corresponding to Amplicons A, B and C were detected in 20 (100%), 19 (95%) and 20 (100%) samples respectively, out of a total of 20 samples. Hence, the analytical limit of detection, defined as the lowest detectable amount of SARS-CoV-2 RNA input at which ≥ 95% of all true positive replicates test positive, was determined to be 100 copies per reaction for Amplicon C, and 250 copies for both Amplicons A and B. Therefore, the overall analytical limit of detection of MassMark was 100 copies per reaction.

**Figure 2:**
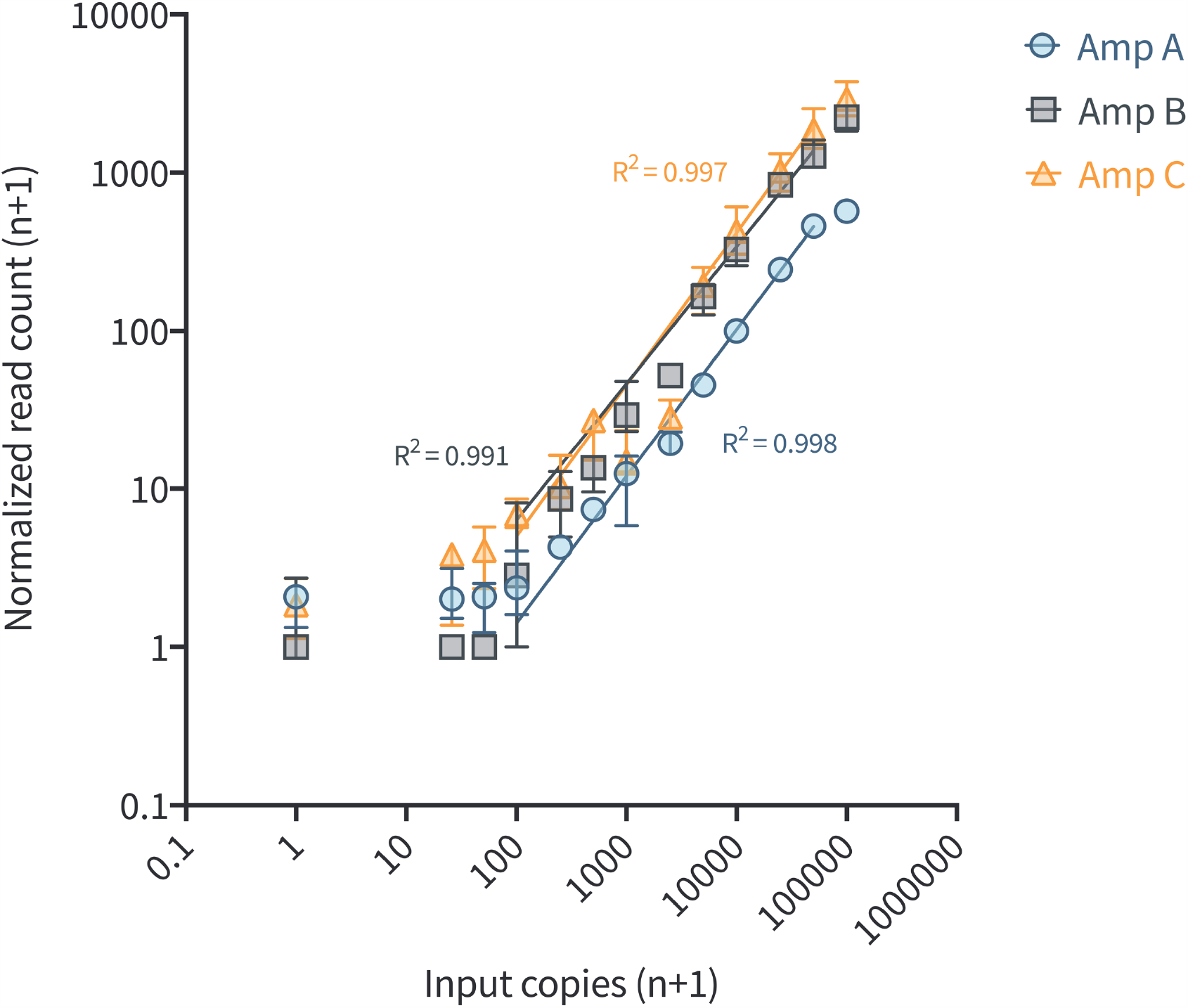
Read counts are correlated to input copy numbers of samples of synthetic SARS-CoV-2 RNA converted to sequencing library using the MassMark method. Median read counts are plotted (n=3). Error bars denote interquartile range. A linear relationship is observed between 100 and 50,000 copies input for amplicons A, B and C.

**Figure 3:**
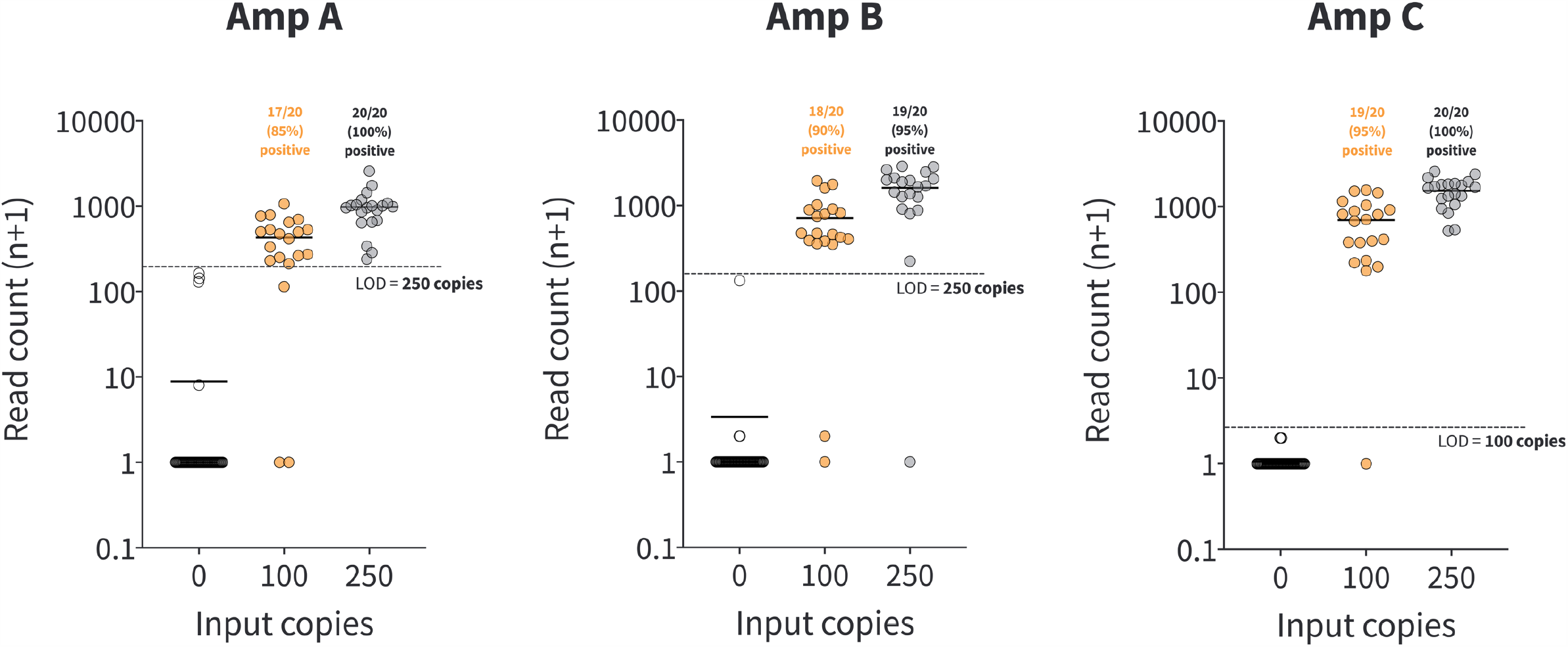
Analytical limit of detection (LOD) for three amplicons A, B and C in MassMark. Samples were run with 0 copies (n=56), 100 copies (n=20) and 250 copies (n=20) of input synthetic SARS-CoV-2 RNA. Solid horizontal lines represent mean read counts. Dotted lines separate read counts obtained for replicates of 0 copies and replicates of copy number representing the LOD of MassMark assay for each of the three amplicons. The LOD differs for each amplicon and is set to 100 copies per reaction for the overall assay.

### Clinical validation

To validate the clinical performance of MassMark, we performed the MassMark protocol on 22 clinical samples which had been orthogonally tested for SARS-CoV-2 with an inhouse RT-PCR Laboratory Developed Test (LDT) utilizing primers and probes from the Centers for Disease Control and Prevention (CDC) 2019-Novel Coronavirus (2019-nCoV) Real-Time RT-PCR Diagnostic Panel^3^ with a target size within 70 bp. The 22 clinical samples comprised of 12 SARS-CoV-2 positive samples with mid to late cycle threshold (Ct) values (range: 22.98-32.72) and 10 SARS-CoV-2 negative samples across three different sample types nasopharyngeal swabs, nasal swabs and saliva) (Table 1). To evaluate the performance of MassMark in a larger background of samples, we also included an additional 415 negative no-template samples. This translates to 12 SARS-CoV-2 positive samples (2.7%) out of a total of 437 samples, which would be in line with the approximately 1-4% prevalence of SARS-CoV-2 infections observed from a number of surveillance^5,7,28^ and mass screening^29^ efforts in places where the virus is actively spreading. Each plate also includes negative controls (0 copies) and positive controls from SARS-CoV-2 RNA at the analytical LOD of each of the three amplicons (100 copies or 250 copies) at fixed well positions per plate (Fig S1). The 22 clinical samples were distributed randomly across five 96-well plates (Fig S1). Positive/negative calls were made based on positive and negative controls included on each plate, according to the interpretation guidelines outlined in Fig S2.

**Table 1:**
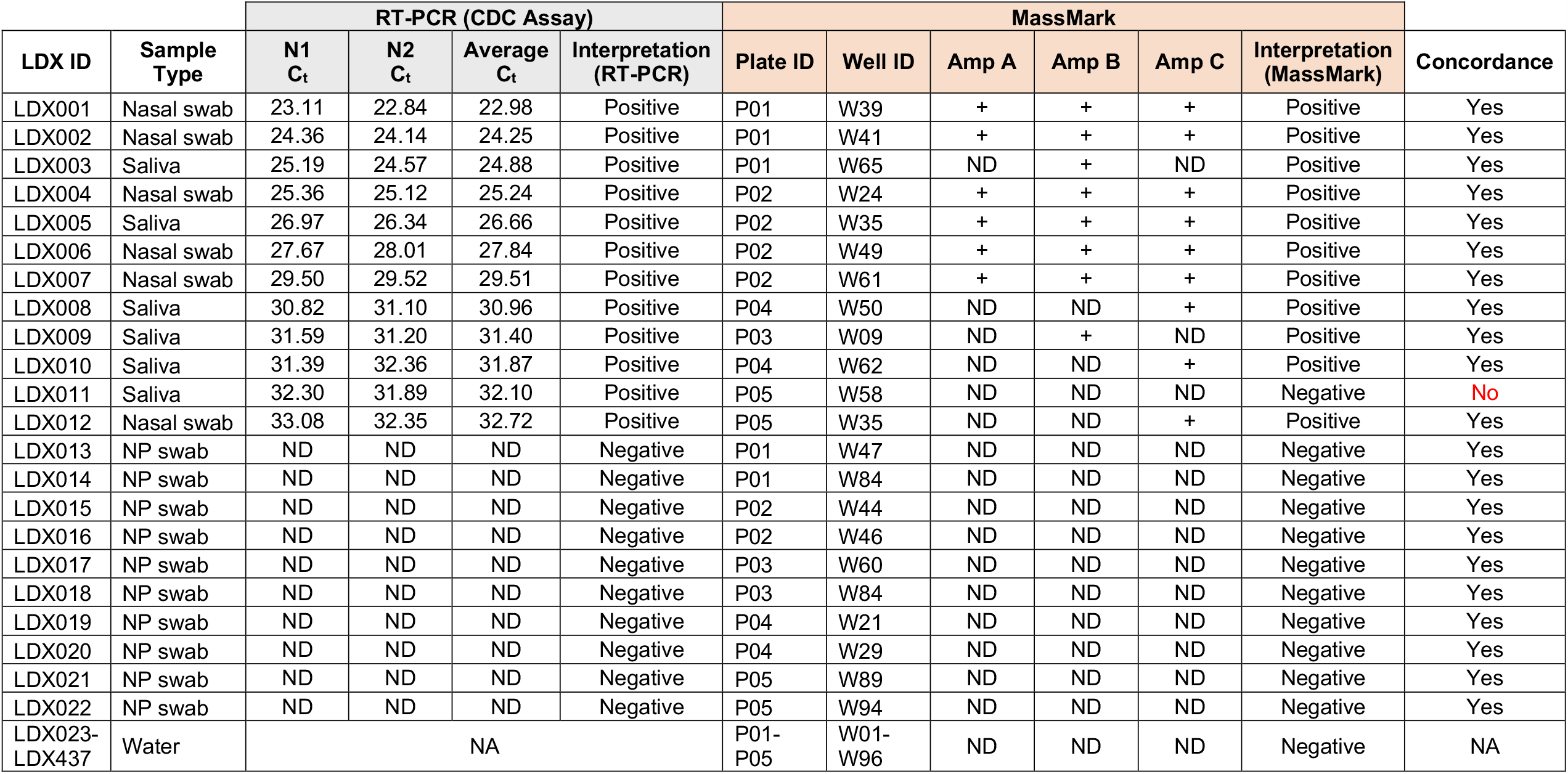
Summary of results for 22 clinical samples tested with MassMark in a sequencing run with 437 total samples.

Out of 12 SARS-CoV-2 positive clinical samples tested, we were able to detect SARS-CoV-2 reads above the detection limit in 11 samples via MassMark (Table 1, Fig 4A). There were no false positives obtained among 10 SARS-CoV-2 negative clinical samples and 415 negative no-template samples (Table 1, Fig 4A). Read counts obtained for positive clinical samples were correlated with the Ct values previously determined by RT-PCR (Fig 4B). The single false negative sample (LDX011) had a late average RT-PCR Ct value of 32.10. Since the RT-PCR assay gives an average Ct value of 34.15 with 25 copies of SARS-CoV-2 RNA input (Table S1), LDX011 would approximately contain less than 100 copies of SARS-CoV-2 RNA, which is below the limit of detection of MassMark. Overall, for the 22 clinical samples tested, the concordance of SARS-CoV-2 detection compared to RT-PCR was high, with one false negative only (Table 2). The sensitivity of MassMark was 91.67% and specificity was 100% (Table 3).

**Table 2:** Concordance between MassMark and RT-PCR for SARS-CoV-2 diagnosis in 22 clinical samples.

|  |  | MassMark |  |
| --- | --- | --- | --- |
|  |  | SARS-CoV-2 Positive | SARS-CoV-2 Negative |
| RT-PCR | SARS-CoV-2 Positive | 11 | 1 |
|  | SARS-CoV-2 Negative | 0 | 10 |

**Table 3:** Performance parameters of MassMark across 22 clinical samples.

|  |  |  |  |
| --- | --- | --- | --- |
| **Sensitivity** | TP/(TP+FN) | 11/(11+1) | 91.67% |
| **Specificity** | TN/(TN+FP) | 10/(10+0) | 100.00% |

**Figure 4:**
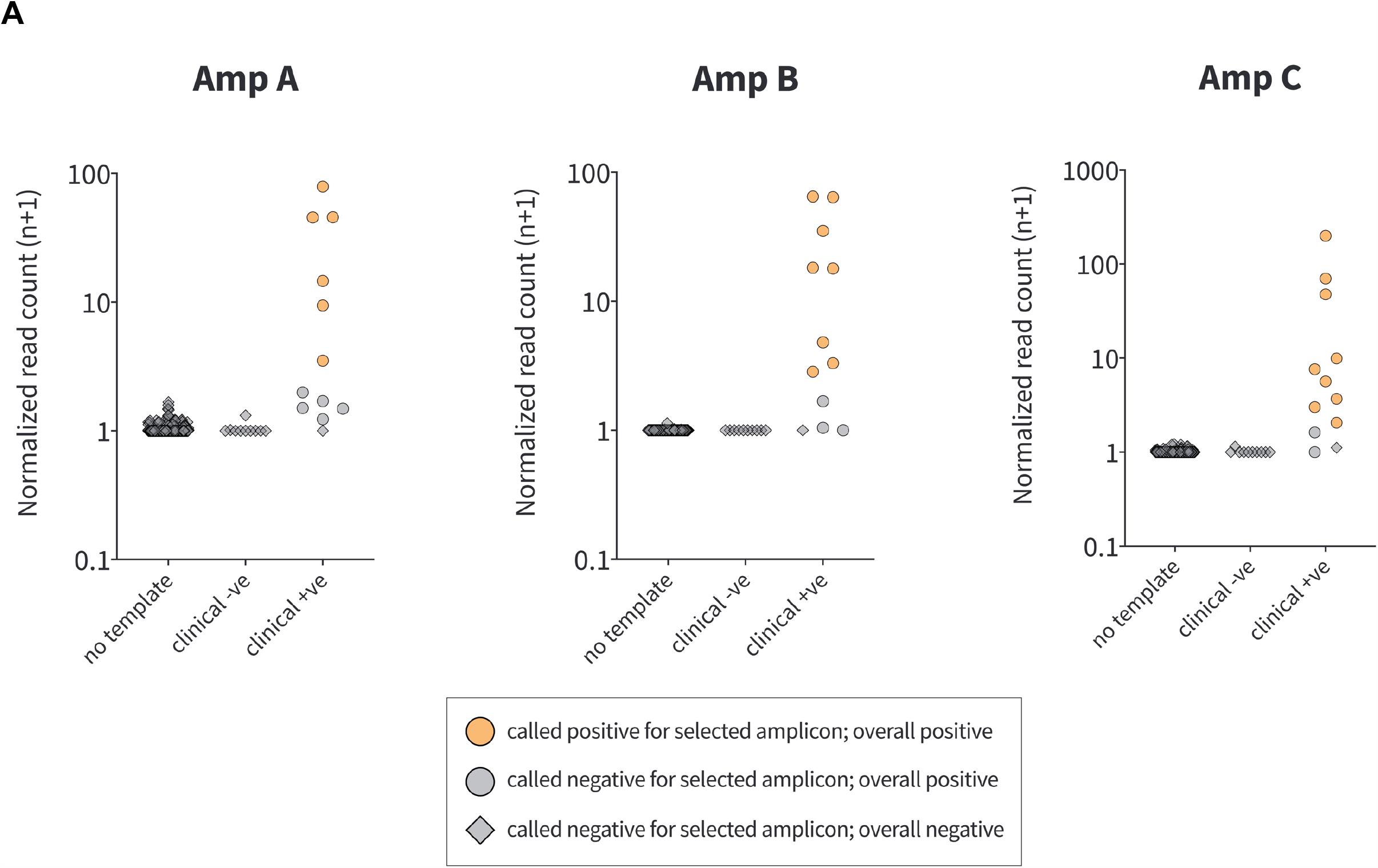
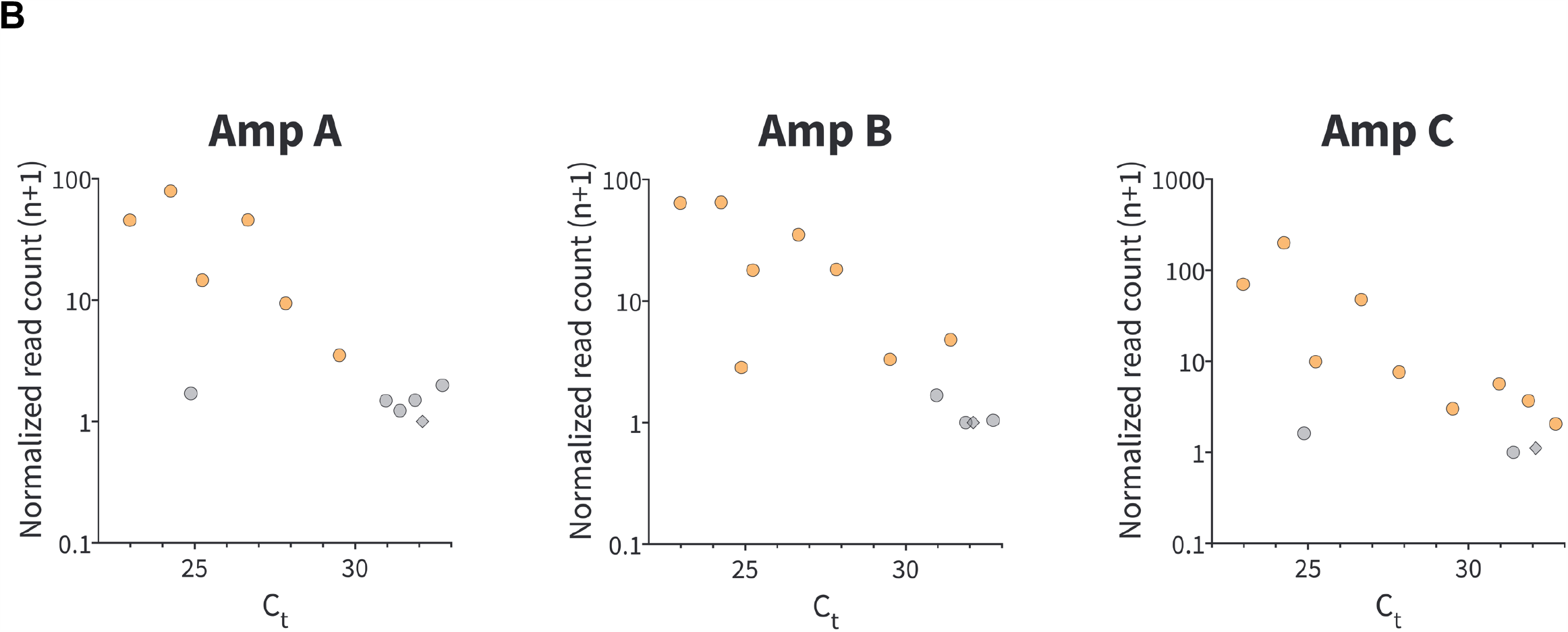
**(A)** Distribution of normalized sequencing read counts for contrived mix of 437 samples, including negative no-template samples (n=415), SARS-CoV-2 negative clinical samples (n=10) and SARS-CoV-2 positive clinical samples (n=12), across all three amplicons A, B and C in MassMark. Raw read counts were normalized to internal plate positive controls. **(B)** Normalized sequencing read counts (MassMark) vs Ct values (RT-PCR) for 12 positive clinical samples. In general, lower read counts are obtained for samples with higher Ct values.

## Discussion

Mass testing has been mooted as the most practical and straightforward way^1,2^ to curb the SARS-CoV-2 pandemic, as it allows health authorities to rapidly identify and isolate infectious individuals, thus limiting the spread of disease and minimizing the need for drastic widespread lockdown measures, which can have negative socioeconomic consequences^15^. Even in places which have successfully controlled the initial wave of SARS-CoV-2 infections, population-scale mass testing has been proven to be helpful in identifying asymptomatic cases^30,31^ that have been missed by symptom-based detection regimes but could still contribute to silent transmission of the virus^18,19^.

With its intrinsic properties of being highly scalable and high throughput, NGS lends itself naturally to mass testing of SARS-CoV-2 as it enables a large amount of data to be generated in a relatively short amount of time, while allowing a high level of specificity to be maintained via mapping of exact sequences. Along these lines, a number of different NGS-based approaches have been proposed for SARS-CoV-2 diagnostics, including LAMP-Seq^23^, DxSeq^24^, REMBRANDT^25^, HiDRA-seq^26^ and Swab-Seq^27^, all of which utilize molecular barcodes for sample identification. Similarly, MassMark identifies patient samples by labelling each sample with a unique, predetermined combination of index sequences, which enables subsequent demultiplexing following sequencing with NGS.

However, unlike other NGS-based approaches, MassMark employs a novel multi-step pooling approach where samples are combined together in consecutive steps, resulting in a single library per workflow with up to 9,216 samples represented. Multiple libraries can be further multiplexed to increase testing capacity per sequencing run. The serial pooling strategy of MassMark reduces handling complexity and conserves resources after every pooling step, thus allowing costs to be kept to a minimum, which is an important and desirable feature in the context of mass screening. Multiplex detection of three specific SARS-CoV-2 amplicons increases detection sensitivity while preserving a high level of specificity. By including multiple amplicons, the chances of encountering false negatives due to *de novo* mutations arising in primer binding sites are reduced, since the detection of a single amplicon is sufficient to call the sample positive. In our study, detection sensitivity is improved by integrating information from all three amplicons, compared to using any single amplicon alone (Table 1).

While RT-PCR tests are typically more sensitive than MassMark, slightly lower sensitivity is an acceptable trade-off for higher throughput in the context of mass screening and surveillance^32^. We show that MassMark is almost as sensitive as RT-PCR for detecting SARS-CoV-2 positive clinical samples with mid to late Ct values (up to Ct 33). Compared to existing methods for SARS-CoV-2 mass screening, MassMark offers a sensitivity that is several orders of magnitude higher than rapid antigen tests^33,34^, at the cost of increased minimum assay turnaround time, though not necessarily overall throughput. The turnaround time for MassMark can be further improved by sequencing only the molecular barcodes needed for sample identification, instead of the whole amplicon. In addition, the MassMark workflow can be further streamlined by investigating its compatibility with direct, extraction-free detection of SARS-CoV-2 from patient samples, which has been extensively studied and validated with RT-PCR assays^35–37^.

In terms of detection specificity, MassMark offers an improvement over existing methods of RT-PCR and rapid antigen tests. Sequenced reads are mapped to the SARS-CoV-2 genome, thus providing further confidence of an accurate call and minimizing the number of false positives. In our study of over 400 samples, no false positives were observed, indicating a high level of specificity, although this would have to be further validated when MassMark is deployed on a larger scale. MassMark also allows genotyping of a limited number of hotspot regions in SARS-CoV-2, with Amplicon B designed to detect the D614G mutation, a major variant in the spike protein of SARS-CoV-2^38^. Should the need arise, other SARS-CoV-2 mutation hotspots can be potentially targeted as well by designing alternative amplicons. Since MassMark is compatible with multiplexing, the method can also be readily extended to target and differentiate other existing pathogens e.g. influenza viruses in the same assay or even novel pathogens in future pandemics.

On its own, MassMark is by no means a complete solution for mass testing of SARS-CoV-2 as it requires specialized infrastructure. However, a ready source of NGS capacity is already available at existing sequencing facilities, which can potentially be repurposed in order for mass testing, which would require millions of tests per country per week^39^, to be feasible. MassMark can be part of a solution for scaling testing capacity at a centralized mass testing facility, and would have the advantage of being easily compatible with automation as well. A single benchtop sequencer has sufficient capacity to sequence > 10,000 MassMark samples in a single run (Table S2); with more sequencers even an average sequencing facility will be able to perform hundreds of thousands of tests in a single day.

In summary, MassMark offers a unique feature set that is complementary to existing SARS-CoV-2 diagnostic methods of RT-PCR and rapid antigen tests, and would be a viable alternative option for scaling SARS-CoV-2 diagnostics especially in the context of mass screening and surveillance, which still have an important role to play in pandemic management while effective vaccines are being manufactured and made widely available.

## Materials and Methods

### Sample Collection and Processing

Clinical specimens including nasopharyngeal swabs, nasal swabs and saliva were collected as part of a study that was evaluated and approved by the Director of Medical Services, Ministry of Health, under Singapore’s Infectious Diseases Act^40^. Viral nucleic acid was extracted from the specimens using Viral Nucleic Acid Extraction Kit II (Geneaid Biotech Ltd., Taiwan) according to the manufacturer’s instructions. The extracted nucleic acid was stored at -20°C. Samples were processed in a College of American Pathologists (CAP) accredited and a Clinical Laboratory Improvement Amendments (CLIA) licensed laboratory (Lucence) and archived extracted RNA from anonymized samples were used.

### RT-PCR Laboratory Developed Test

Real-time reverse transcription-polymerase chain reaction (RT-PCR) is employed to screen for presence of SARS-CoV-2 RNA using N1 and N2 markers as specified in the CDC 2019-nCoV Real-Time RT-PCR Diagnostic Panel^3^. 7.5 μl of extracted RNA was mixed with 10 μl Luna Universal One-Step Reaction Mix (2x) (New England Biolabs, Ipswich, MA, USA), 1 μl Luna WarmStart RT Enzyme Mix (20x) (New England Biolabs), 1.5 μl N1 primers+probe mix (Integrated DNA Technologies, Coralville, IA, USA) or 1.5 μl N2 primers/probe mix (Integrated DNA Technologies) in a 20 μl reaction. Reactions were carried out in a CFX96 Touch Real-Time PCR Detection System (Bio-Rad, Hercules, CA, USA), with reverse transcription at 55°C for 10 min, then an initial denaturation step at 95°C for 1 min, followed by 45 cycles of 95°C denaturation for 10s and 60°C extension for 30s. Fluorescence intensity is captured at the end of every extension cycle, and a cycle threshold (Ct) value is calculated using the Bio-Rad CFX Manager 3.1 software. Limit of detection analysis for the RT-PCR LDT is shown in Table S1.

### MassMark Primer Design

Primers were designed against the SARS-CoV-2 Reference Genome (NC_045512.2). We utilised the Nextstrain^41^ and GISAID^42^ databases of SARS-CoV-2 sequences to identify regions of low mutational entropy and minimize the chances of encountering mutations within the primer binding sites. Three pairs of primers were designed to capture sequences corresponding to three different target regions of SARS-CoV-2. Two of these amplicons (Amplicons A and B) are located within the S gene of SARS-CoV-2, while the remaining amplicon (Amplicon C) lies in the N gene.

### Specificity/Exclusivity Testing: In Silico Analysis

BLASTn analysis queries of the primer pairs used in MassMark were performed against a public domain nucleotide sequence database (updated on 07/12/2020) consisting of GenBank+EMBL+DDBJ+PDB+RefSeq sequences, but excluding EST, STS, GSS, WGS, TSA, patent sequences as well as phase 0, 1, and 2 HTGS sequences and sequences longer than 100Mb. The search parameters automatically adjust for short input sequences, with match and mismatch scores set at 1 and -3 respectively, and gap costs for creation and extension at 5 and 2 respectively.

#### Amplicon A

Forward primer showed high sequence homology with Pangolin coronavirus and Bat coronavirus genomes, while reverse primer showed high sequence homology with Bat coronavirus genome. Taking into account both primers, there are no significant homologies with human genome, human pathogens or human microflora that would predict potential false positive results.

#### Amplicon B

Both forward and reverse primers did not show significant homology with other coronaviruses. Taking into account both primers, there is no prediction of potential false positive results with human genome, human pathogens or human microflora.

#### Amplicon C

Forward primer showed high sequence homology with Bat coronavirus genome, while reverse primer showed high sequence homology with SARS coronavirus, Pangolin coronavirus and Bat coronavirus genomes. Taking into account both primers, there are no significant homologies with human genome, human pathogens or human microflora that would predict potential false positive results.

In summary, the MassMark assay, which is designed for the specific detection of SARS-CoV-2, showed no significant combined homologies with human genome, human pathogens or human microflora that would predict potential false positive results.

### Reverse Transcription with Indexing (or Barcoding)

Reverse transcription (RT) was performed using the High-Capacity cDNA Reverse Transcription Kit (Applied Biosystems, Waltham, MA, USA) according to the manufacturer’s instructions. Briefly, 10 μl of extracted RNA was mixed with 2 μl 10X RT Buffer, 0.8 μl 100mM dNTPs, 1 μl Multiscribe Reverse Transcriptase, 1 μl RNase inhibitor, 2 μl 1μM barcoding (well) RT primers (Integrated DNA Technologies), 1 μl 4.15fM spike-in control (Integrated DNA Technologies) and nuclease-free water in a 20 μl reaction. Each reaction consists of a set of three barcoding RT primers, with one RT primer for each of the three target amplicons (Amplicons A, B, C). Reactions were carried out in a C1000 Touch Thermal Cycler (Bio-Rad) at 25°C for 10 min, followed by 37°C for 120 min and finally 85°C for 5 min. First strand cDNA products were immediately used in the subsequent steps or stored at -20°C.

### Internal and External Reaction Controls

The spike-in control, a synthetic RNA with primer binding sites for reverse transcription and PCR primers corresponding to Amplicon A, was included in every reverse transcription reaction at 2,500 copies per well as an internal positive control. Other than the primer binding sites, the spike-in control contains a pseudo sequence which allows it to be distinguished from Amplicon A reads originating from actual SARS-CoV-2 viral RNA.

Four external positive and four external negative controls were also included on every plate in fixed positions. The four external positive controls comprised of synthetic SARS-CoV-2 RNA (Twist Bioscience, San Francisco, CA, USA) at 10,000, 1,000, 250 and 100 copies per well.

The negative, no template controls comprised of nuclease-free water instead of extracted sample RNA.

### PCR for Amplicon Generation with Indexing (or Barcoding)

A maximum of 96 first strand cDNA samples per plate were pooled together and concentrated in an Amicon Ultra-2 Centrifugal Filter (Merck, Darmstadt, Germany), followed by a round of purification with AMPure XP beads (Beckman Coulter, Brea, CA, USA). A limited, single-cycle amplification of the pooled cDNA products using a single barcoding primer for each of the three amplicons was performed to generate double-stranded cDNA products. Briefly, 25 μl of purified first strand cDNA was mixed with 5 μl 5X SuperFi II Buffer, 1 μl 10mM dNTPs, 1 μl Platinum SuperFi II DNA (Invitrogen, Waltham, MA, USA), 1 μl 5μM barcoding (plate) forward primers (Integrated DNA Technologies) and nuclease-free water in a 50 μl reaction. Reactions were carried out in a C1000 Touch Thermal Cycler at 98°C for 90 s, 60°C for 6 min and finally 72°C for 6 min. Double-stranded cDNA products were immediately used in the subsequent steps or stored at -20°C.

### Product Amplification

A maximum of 96 double-stranded cDNA samples were pooled together and concentrated in an Amicon Ultra-2 Centrifugal Filter (Merck, Darmstadt, Germany). The concentrated product was subjected to treatment with 50 units of Thermolabile Exonuclease I (New England Biolabs) in 1x NEBuffer 3.1 in a 50 μl reaction at 37°C for 10 min and then 80°C for 1 min, followed by a round of purification with AMPure XP beads. To amplify products and add appropriate adaptors for compatibility with Illumina sequencing, PCR was performed with KAPA HiFi HotStart ReadyMix according to the manufacturer’s instructions. Briefly, 23 μl of purified double-stranded cDNA was mixed with 25 μl 2X KAPA HiFi HotStart ReadyMix, 1 μl 20μM indexed P5 primer (Integrated DNA Technologies) and 1 μl 20μM indexed P7 primer (Integrated DNA Technologies). Reactions were carried out in a C1000 Touch Thermal Cycler with an initial denaturation step at 98°C for 45 s, followed by 24 cycles of denaturation (98°C, 15 s), annealing (60°C, 30 s) and extension (72°C, 30 s). This was then finished with a final extension step at 72°C for 1 minute. The amplified libraries underwent two rounds of purification with AMPure XP beads. The final purified libraries were visualized with a High Sensitivity D1000 Screentape (Agilent, Santa Clara, CA, USA) on a 4200 Tapestation (Agilent). Libraries were quantified using a KAPA Library Quantification Kit (Roche, Basel, Switzerland) on a CFX96 Touch Real-Time PCR Detection System, before sequencing on a MiniSeq (Illumina, San Diego, CA, USA) or NextSeq 550 (Illumina) depending on the sample load (Table S2). 35,000 reads are allocated per individual sample.

### Indexes

Well and plate indexes are all 10-bp long and are predetermined to have a minimum edit distance of 3 from every other index. To achieve multiplexing of 9,216 samples, 96 well indexes and 96 plate indexes are required.

### Sequence Deconvolution and Sample Identification

Demultiplexed FASTQ files were analyzed for expected well and plate indexes that occur at the beginning and end of reads and correspond to single specimens. Reads were matched to targeted SARS-CoV-2 genome and enumerated for each well and plate index combination available in the predefined index library. Reads were also matched to the spike-in control sequence with same index combinations and enumerated. Specimens were positive for SARS-CoV-2 if they were represented by well and plate index combinations, among the sequencing reads. No corresponding well and plate index combination is expected for specimens with no SARS-CoV-2.

## Data Availability

The datasets generated during and/or analysed during the current study are available from the corresponding author on reasonable request.

## Acknowledgements

We would like to acknowledge Ziyi Wan and Ramandeep Virk for their support with samples and sequence analysis for primer design, and the medical laboratory technologists at Lucence service laboratory.

## Supplementary Figures and Tables

**Figure S1:**
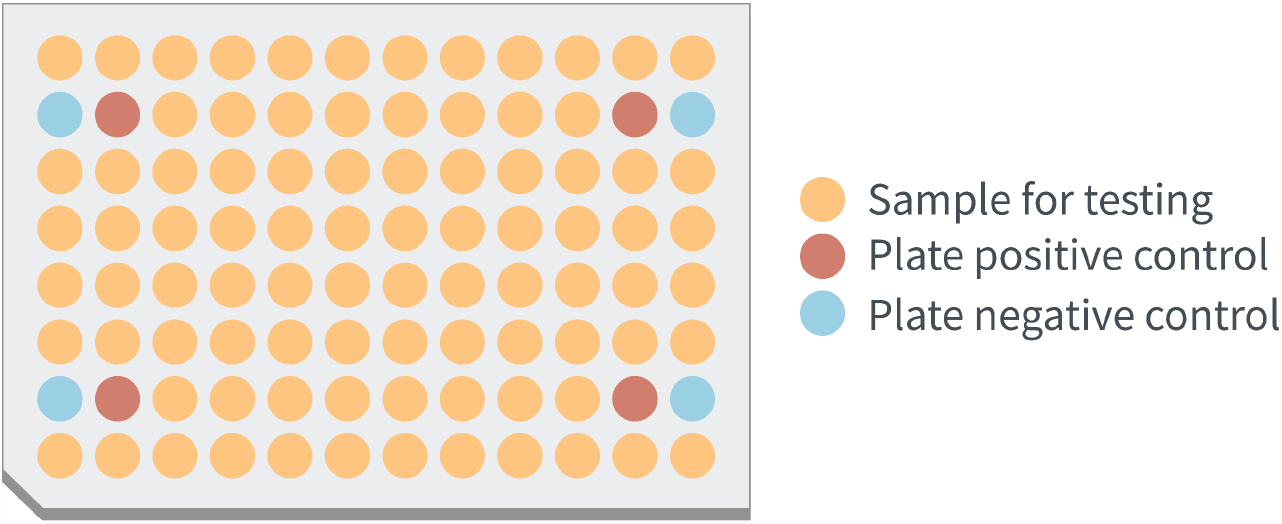
Sample 96-well plate layout for MassMark

**Figure S2:**
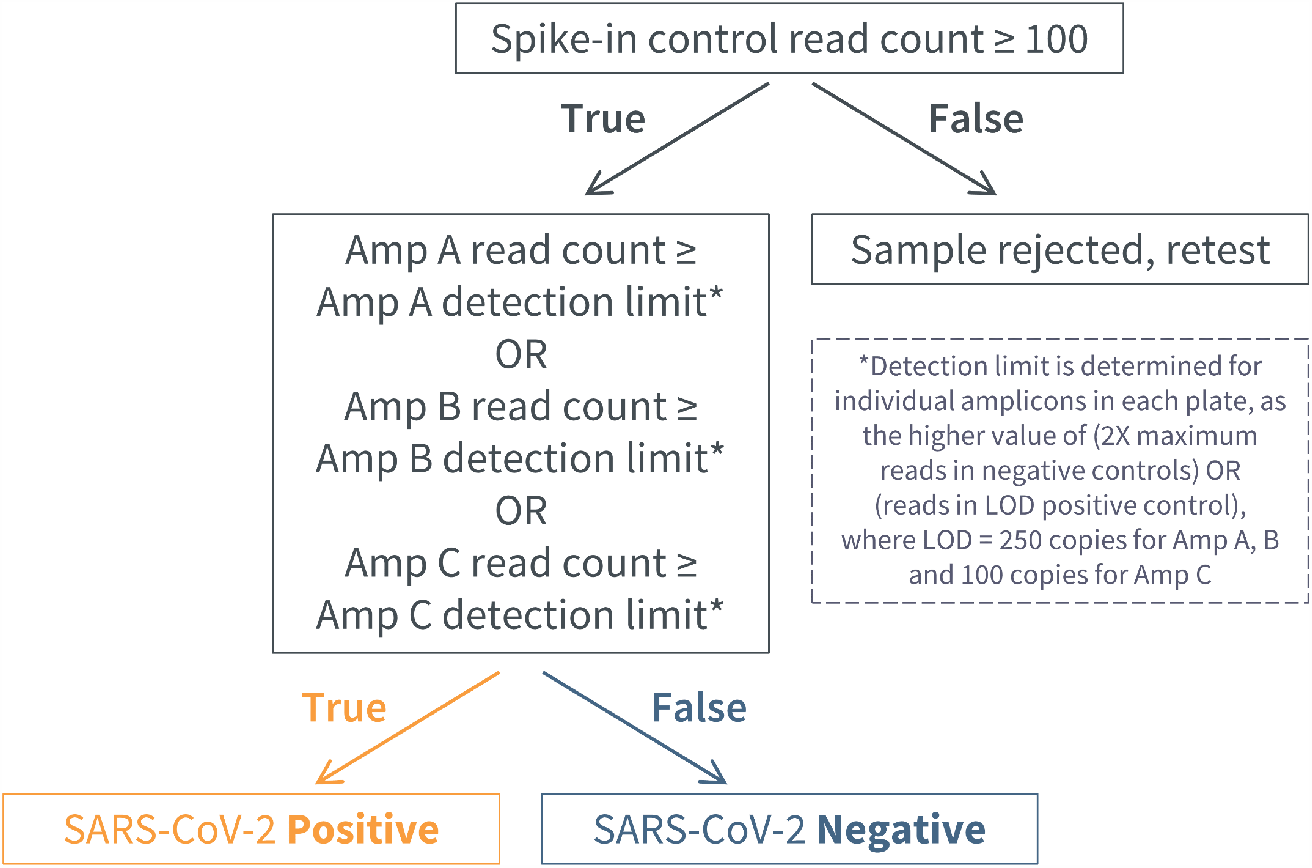
Flowchart for interpreting MassMark results for SARS-CoV-2 detection

**Table S1:** Limit of detection of RT-PCR assay based on CDC 2019-nCoV Real-time RT-PCR Diagnostic Panel.

| **Targets** | **2019-nCoV_N1** | **2019-nCoV_N2** |
| --- | --- | --- |
| **RNA copies per reaction** | 25 | 25 |
| **n Positives/Total** | 19/20 | 20/20 |
| **Mean C<sub>t</sub>** | 34.28 | 34.01 |
| **Standard Deviation (C<sub>t</sub>)** | 0.94 | 0.95 |

**Table S2:** Comparison of high-throughput sequencer capacity for MassMark samples.

| **Sequencing Platform**<br>**(Illumina)** | **Maximum number of**<br>**MassMark samples per run** |
| --- | --- |
| MiniSeq (High-Output Kit) | 700 |
| NextSeq 550 (High-Output Kit) | 11,500 |
| NovaSeq 6000 (S1 Flow Cell) | 46,000 |

